# Treatment interruption is a risk factor for sickness presenteeism: a large-scale cross-sectional study during the COVID-19 pandemic

**DOI:** 10.1101/2021.08.14.21261996

**Authors:** Makoto Okawara, Tomohiro Ishimaru, Seiichiro Tateishi, Ayako Hino, Mayumi Tsuji, Akira Ogami, Tomohisa Nagata, Shinya Matsuda, Yoshihisa Fujino, for the CORoNaWork project

## Abstract

**Objectives:** This study examined the relationship between interruption to routine medical care during the coronavirus disease 2019 pandemic and sickness presenteeism among workers in Japan.

**Methods:** A cross-sectional study using data obtained from an internet monitor questionnaire was conducted. Interruption to medical care was defined based on the response “I have not been able to go to the hospital or receive treatment as scheduled.” The fraction of sickness presenteeism days in the past 30 days was employed as the primary outcome. A fractional logit model was used for analysis to treat bounded data.

**Results:** Of the 27,036 participants, 17,526 (65%) were workers who did not require routine medical care, 8,451 (31%) were using medical care as scheduled, and 1,059 (4%) experienced interrupted medical care. The adjusted odds ratio (aOR) of sickness presenteeism was significantly higher among workers who experienced interrupted medical care (3.44; 95% confidence interval [CI]: 3.04–3.89) than those who did not require routine medical care. In terms of symptoms, the highest aOR was observed among workers with mental health symptoms (aOR: 5.59, 95%CI: 5.04–6.20).

**Conclusions:** This study suggests the importance of continuing necessary treatment during a pandemic to prevent presenteeism.

## Introduction

Sickness presenteeism is an increasingly important issue in occupational health. Aronsson defined sickness presenteeism as “people, despite complaints and ill health that should prompt rest and absence from work, still turning up at their jobs”^1^. Sickness presenteeism is the result of a choice made by a worker with ill-health, disease, or capacity loss between sickness presenteeism and sickness absence^2^. This decision is influenced by the individual’s personality, values, economic status, workplace “demands for presence” and support for adaptation, and national culture and employment customs^2^. Evidence suggests that sickness presenteeism can lead to sickness absence and future worsening of physical and mental health conditions^3–9^. In addition, the impact of working while ill on productivity is also gaining attention, especially in North America^10,11^. A variety of diseases and health conditions have been found to be associated with sickness presenteeism, suggesting the importance of managing disease and maintaining good condition^4,12^.

Under the coronavirus disease 2019 (COVID-19) pandemic, there is concern that both organizational and individual factors will increase sickness presenteeism above that observed under normal conditions^13–15^. Increased workload on workers can lead to a negative work culture around taking sick leave, such as where workers who choose to work while ill are valued for their loyalty to the company and motivation to work, thus promoting sickness presenteeism^16^. Examples of individual factors that may increase sickness presenteeism include worsening economic situations and job insecurity; increased telecommuting, which can make it easier for workers to work even while sick; the impact of the pandemic on anxiety and mental health; worsening of health conditions and diseases due to lifestyle changes; and worsening of chronic diseases due to the inability to access medical resources. All of these factors are expected to lead to an increase in sickness presenteeism.

Interruption to medical care is an important problem in the COVID-19 pandemic. Access to necessary routine medical care and medical resources is reportedly being affected in many countries around the world^17–19^. In Japan, there is data showing that the number of prescriptions issued has decreased^20^. Interruption to medical care can adversely affect management of chronic diseases and delay the detection and treatment of new diseases^22^. In fact, excess deaths unrelated to COVID-19 have been reported^23^. Thus, interruption to medical care during the COVID-19 pandemic may lead to worsening of non-COVID-19 diseases and health conditions.

During the COVID-19 pandemic, the number of people working while ill may increase as a result of worsening health conditions arising from treatment interruptions and delays. This may result in an increase in sickness presenteeism. However, few studies have examined the effect of medical care interruption on sickness presenteeism during the COVID-19 pandemic. We hypothesized that sickness presenteeism has increased among workers who experienced interruptions to their medical care during the COVID-19 pandemic. The purpose of this study was to examine the association between medical care interruption and sickness presenteeism in Japanese workers during the COVID-19 pandemic.

## Materials and Methods

We performed a cross-sectional study based on baseline survey data obtained in the Collaborative Online Research on the Novel-Coronavirus and Work (CORoNaWork) project, a prospective cohort study that performed a questionnaire-based survey of Internet monitors to determine the effect of the COVID-19 pandemic on workers’ health. Details of the study protocol are published elsewhere^23^. The practical aspects of the survey, namely recruitment, data sampling, initial data clean-up, and management of respondents’ user identifications (IDs) for tracking cohort data, were conducted by Cross Marketing Inc. (Tokyo, Japan), which has 4.7 million registered monitors. Before completing the online survey, participants read a description of the survey’s aims and details about the handling of their information. Only participants who agreed with the contents of the description were allowed to participate. Participants’ user IDs were stored by the survey company. To participate in surveys offered by the survey company, users had to register in advance. Once registered, participants were given a user ID to use when completing online surveys. Thus, all surveys were conducted anonymously. The survey company assigned each respondent a unique number that would be used only within this study to merge the data from the first and subsequent surveys, based on the user ID within the survey company. The user ID is not provided to the researcher, but the unique number enabled the researcher to merge the data and create the cohort data. Participants’ personal data were anonymized prior to receipt by the researchers, and were protected based on the survey company’s privacy policy. This study was approved by the Ethics Committee of the University of Occupational and Environmental Health, Japan (Approval No. R2-079 and R3-006).

The baseline survey was conducted from December 22 to 26, 2020. A total of 605,381 registered monitors were emailed invitations to participate. Sampling was designed such that sex and occupation (office and non-office workers) were approximately equal among the five regions of residence. Specifically, we predetermined a total of 20 units comprising five regions of residence, two sexes, and two occupations. Sampling was continued until each unit reached 1500 responses plus a margin of 10%, after which the unit was closed to participation. We devised the five regions of residence according to infection and geographic status: first, Japan’s 47 prefectures were divided into four categories based on the cumulative COVID-19 infection rate; second, the category with the highest infection rate was further divided into Kanto and non-Kanto regions. A total of 55,045 participants answered the initial screening questions, of whom 33,302 matched the survey’s criteria (worker status, region of residence, sex, and age). Participants answered one questionnaire item per page, with the overall questionnaire containing 55 pages. Participants could review and change their responses using the back button. Participants who provided fraudulent responses (n=6,266) according to the survey company or a predefined definition of a fraudulent response were excluded. Fraudulent responses included an unusually short response time (below 6 minutes), unusually short height (below 140 cm), unusually low weight (below 30 kg), varying answers to similar questions in the survey (e.g., varying answers to questions about marital status or area of residence), and incorrect answers to tiered questions used to identify inappropriate responses (e.g., choose the third highest number from the following five numbers). After exclusion, responses from 27,036 participants aged from 20 and 65 years who indicated they were working when completing the survey were ultimately included in the analysis.

### Assessment of treatment status

We assessed the presence of disease and use of medical care using the following question: “Do you have a disease that requires regular visits to the hospital or treatment?” Responses were “I do not have any such disease”; “I am able to go to the hospital or receive treatment as scheduled”; “I have not been able to go to the hospital or receive treatment as scheduled.”

Those who answered “I do not have any such disease” were defined as workers who did not require routine medical care, and thus did not have any disease that requires hospital visits or treatment. Those who answered “I am able to go to the hospital or receive treatment as scheduled” were defined as workers who used medical care. Those who answered “I have not been able to go to the hospital or receive treatment as scheduled” were defined as workers who experienced interrupted medical care.

### Assessment of sickness presenteeism and other covariates

Respondents’ number of sickness presenteeism days was ascertained based on the following question and used as the primary outcome: “In the last 30 days, how many days have you worked (including work from home) despite feeling that you really should have taken sick leave due to your state of health?”^2^ According to a previous study, the target period for sickness presenteeism can range from four weeks to 12 months.^24^ We chose a target period of 30 days to reduce the effects of recall bias and changes in the COVID-19 pandemic situation in Japan.

Socioeconomic and work-related factors included sex, age, job type (mainly desk work, mainly interpersonal communication, mainly physical work), marital status (married, divorced/deceased, never married), equivalent income (household income divided by the square root of the number of people in the household), education (junior high school, high school, vocational school/junior college/college of technology, university/graduate school), frequency of working from home (at least one day a month, at least one day a week, at least two days a week, at least four days a week), company size (total number of employees in the respondent’s main place of work [1 for self-employed]), presence of a policy by the employer requesting that employees refrain from attending work while ill (yes or no), and the number of days worked per week.

The cumulative infection rate of COVID-19 in the province of residence was employed as a community-level variable.

To control for potential confounders, we also asked participants to indicate their main symptoms using the following question: “Which of the following conditions or body parts give you the most trouble during your work?” The options were “No problem”; “pain”; “movement”; “tightness, loss of energy, appetite, fever, dizziness, or feeling poor”; “toileting or elimination”; “mental health”; “skin, hair, or beauty”; “sleep”; “eyes”; “nose”; “ears”; and “other.”

### Statistical analysis

Age was treated as a continuous variable and reported as mean and standard deviation (SD). The number of sickness presenteeism days was treated as a discrete variable and converted to a fractional response variable by dividing by the maximum value of 30 days. Categorical variables were reported as number and percentage. Equivalent income was categorized into quartiles.

We compared the results of Poisson regression, Zero-inflated Poisson regression (ZIP), negative binomial regression, Zero-inflated Negative Binomial regression (ZINB), and fractional logistic regression as statistical models. To handle data with excess zeros, which indicates a population at low risk of sickness presenteeism, we considered a zero-inflated model^24^. Fractional logistic regression^26^ was also considered because the maximum possible number of days of sickness presenteeism was 30, allowing the data to be treated as bounded data. As a measure of model fitness, we compared the Akaike’s Information Criterion (AIC), and ultimately adopted the fractional logit model.

Fractional logistic regression analysis was conducted with the fraction of sickness presenteeism days in last 30 days (0 indicates 0 days of sickness presenteeism; 1 indicates 30 days of sickness presenteeism) as the dependent variable and the respondents’ category of treatment status as an independent variable.

We adjusted for the following potential confounders: sex, age, job type, marital status, equivalent income, education, frequency of working from home, company size, cumulative infection rate by prefecture, main symptoms, presence of a policy by the employer requesting that employees refrain from attending work while ill, and the number of days worked per week.

In further analysis, we estimated the margins of sickness presenteeism days for each treatment status and symptom. First, we used the same statistical model as that in the main analysis. Second, we calculated the predictive margins of sickness presenteeism days, substituting measured values for other covariates, dividing the data into 36 groups (3 treatment statuses and 12 symptoms)^27^. In the fractional logit model, because predictive margins were calculated as fractions, we multiplied margins and standard errors by 30 to obtain predictive margins for sickness presenteeism days in the last 30 days. Preliminarily, we confirmed the simple main effects for each treatment status compared to workers who did not require routine medical care by adding the interaction term between treatment status and symptoms to the model used for the main analysis. For all analyses, the Bonferroni method was used to adjust for multiple comparisons.

All comparisons were performed in Stata (Stata Statistical Software: Release 16; StataCorp LLC, TX, USA), with *p*<0.05 indicating statistical significance.

## Results

The demographic and sociological characteristics of the analyzed population are shown in Table 1. A total of 13,814 (51%) were men, and the mean age was 47.0 years (SD: 10.5). Of the total population, 17,526 (65%) were workers who did not require routine medical care, 8,451 (31%) were using medical care as scheduled, and 1,059 (4%) experienced interrupted medical care. The distribution of sickness presenteeism is shown for the three treatment statuses in a histogram in Figure 1. While the majority of respondents reported zero days of sickness presenteeism, a large number also selected the maximum of 30 days. There were also small clusters at 5, 10, and 20 days, which may be due to digit preference.

**Table 1.**
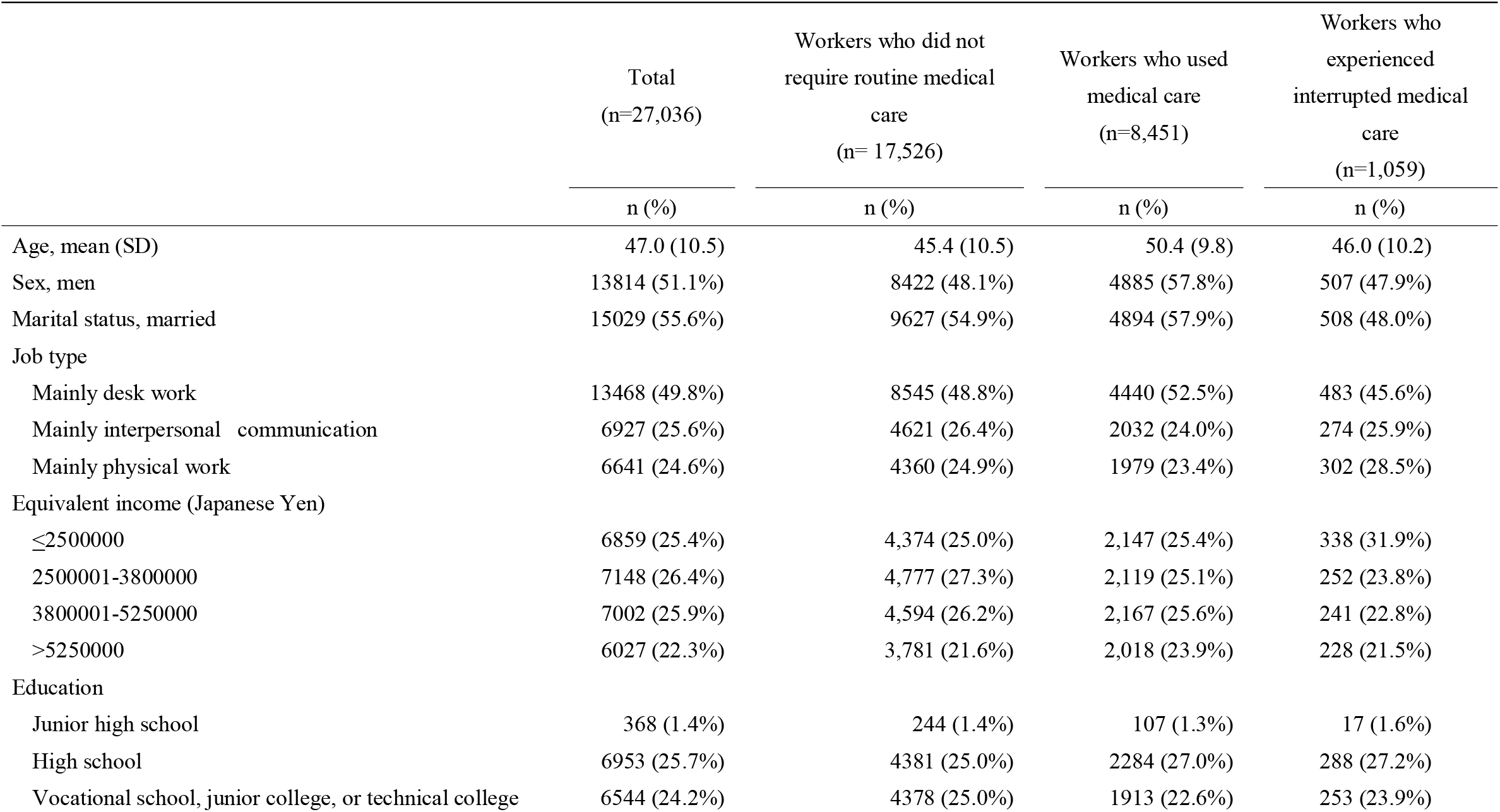
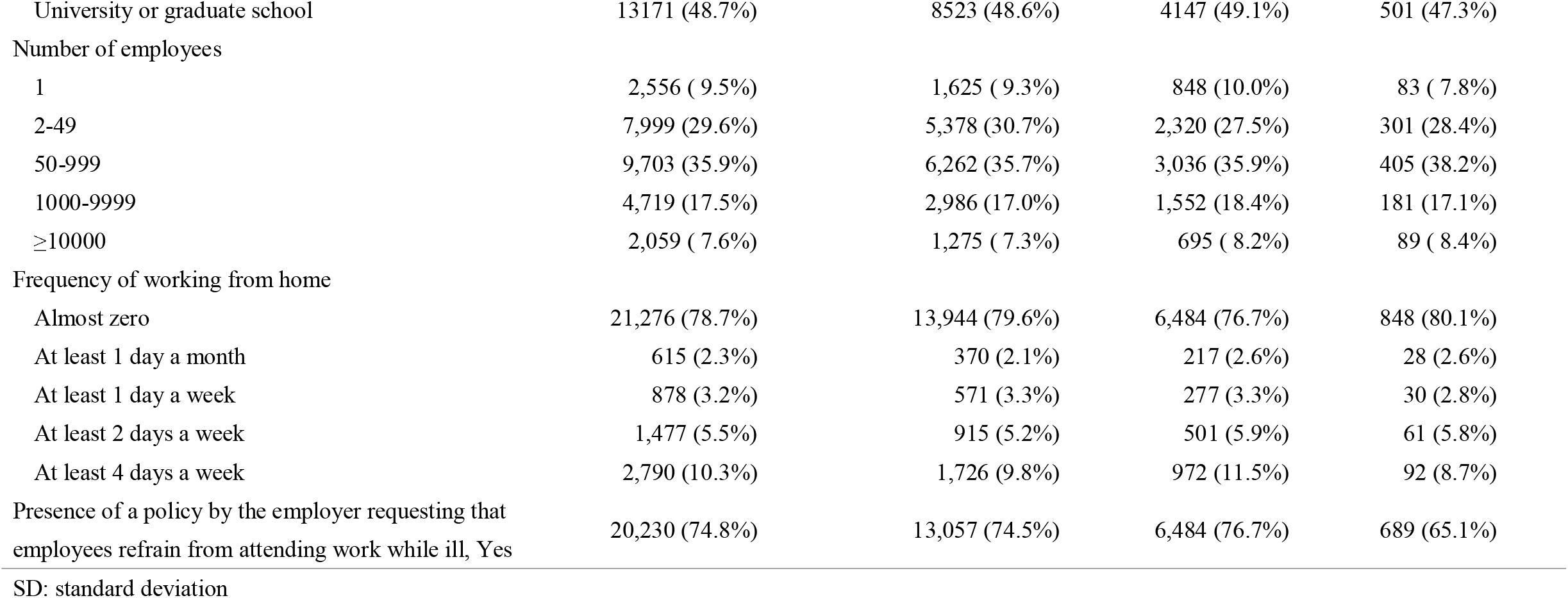
Basic characteristics of the study subjects

**Figure 1.**
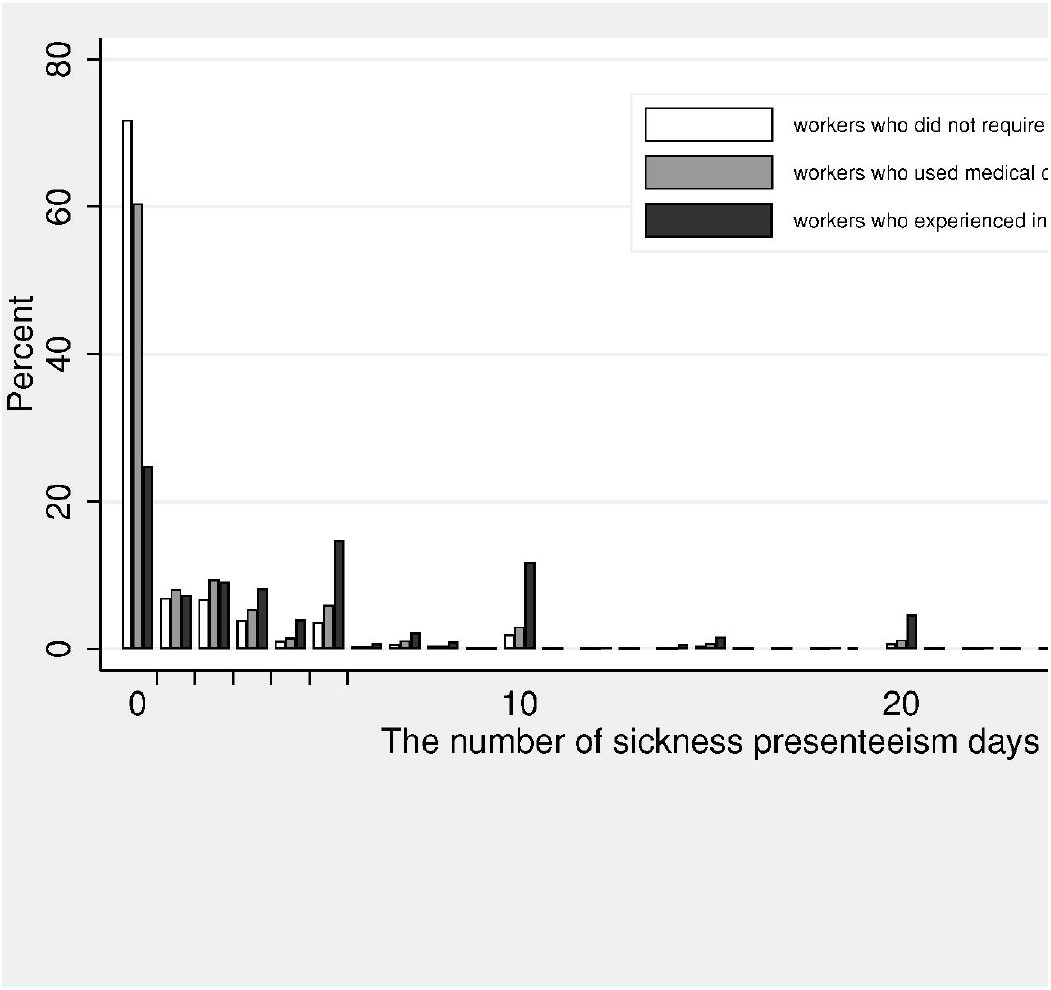
Number of days of sickness presenteeism among workers with each treatment status

The association between treatment status and the fraction of sickness presenteeism days is shown in Table 2. There was a significant association between the fraction of sickness presenteeism days and treatment status. After adjusting for other covariates in the multivariate model, the odds ratio (aOR) of sickness presenteeism days was significantly higher among workers who used medical care (aOR: 1.36, 95%CI: 1.26–1.46, *p*<0.001) and workers who experienced interrupted medical care (aOR: 3.28, 95%CI: 2.93–3.67, *p*<0.001) compared to workers who did not require routine medical care.

**Table 2.** Association between treatment status and sickness presenteeism

| Treatment status | Univariate |  |  |  | Multivariate* |  |  |  |
| --- | --- | --- | --- | --- | --- | --- | --- | --- |
|  | OR | 95% CI |  | *p* | Adjusted OR | 95% CI |  | *p* |
| Workers who did not require routine medical care | Reference |  |  |  | Reference |  |  |  |
| Workers who used medical care | 1.57 | 1.47 | 1.69 | <0.001 | 1.36 | 1.26 | 1.46 | <0.001 |
| Workers who experienced interrupted medical care | 5.75 | 5.16 | 6.40 | <0.001 | 3.28 | 2.93 | 3.67 | <0.001 |

\*Adjusted for sex, age, marital status, equivalent income, education, company size, job type, frequency of working from home, number of days worked per week, presence of a policy by the employer requesting that employees refrain from attending work while ill, cumulative infection rate for COVID-19 in the region of residence, and main symptom.
OR: odds ratio, CI: confidence interval

The association between participants’ main symptoms and the fraction of sickness presenteeism days is shown in Table 3. There were significant associations between the fraction of sickness presenteeism days and some symptoms using the model presented in Table 2. The highest OR of sickness presenteeism days was observed for mental health symptoms (aOR: 5.35, 95%CI: 4.85–5.91, *p*<0.001).

**Table 3.**
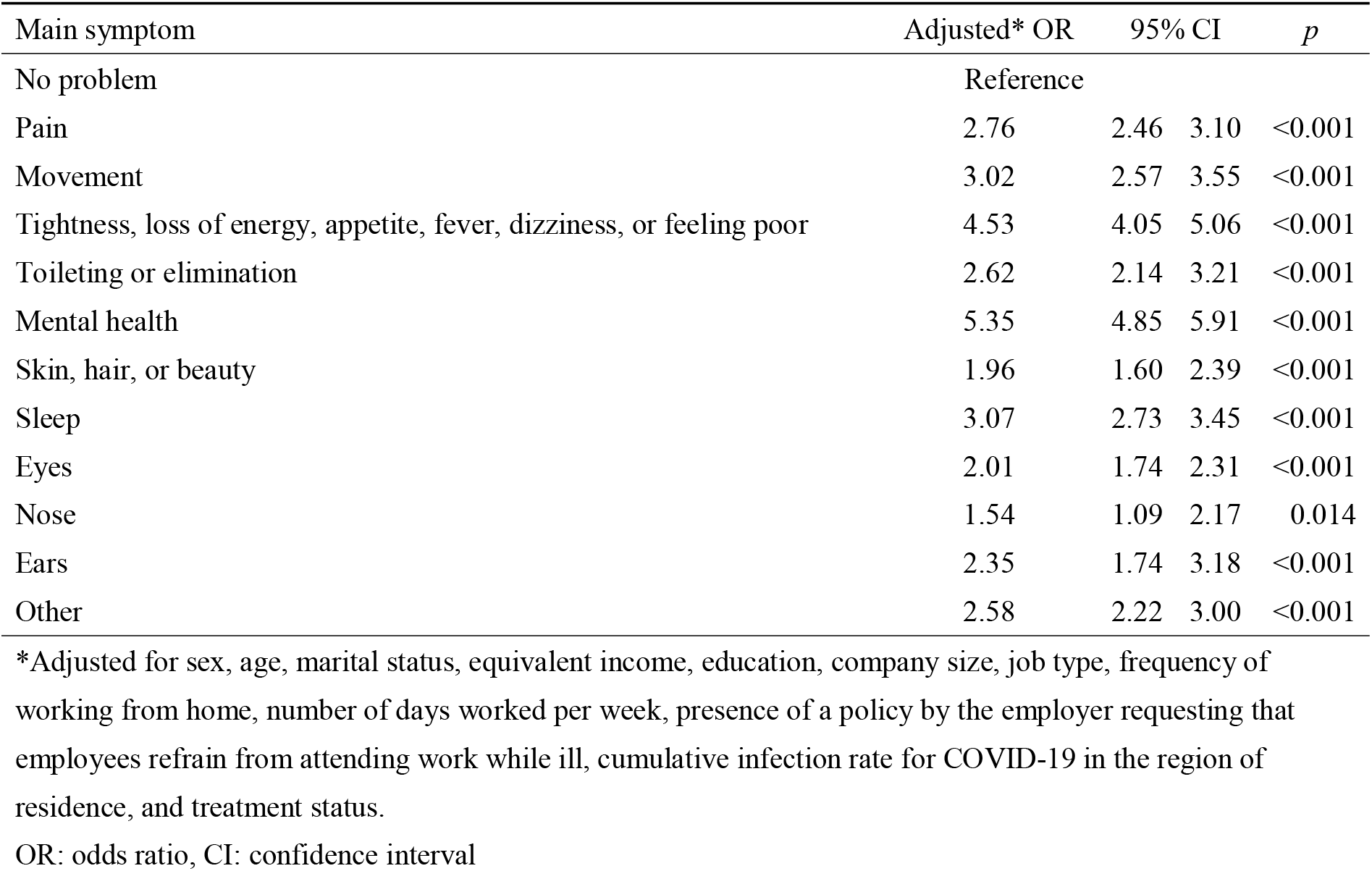
Association between main symptoms and sickness presenteeism

| Main symptom | Adjusted* OR | 95% CI |  | *p* |
| --- | --- | --- | --- | --- |
| No problem | Reference |  |  |  |
| Pain | 2.76 | 2.46 | 3.10 | <0.001 |
| Movement | 3.02 | 2.57 | 3.55 | <0.001 |
| Tightness, loss of energy, appetite, fever, dizziness, or feeling poor | 4.53 | 4.05 | 5.06 | <0.001 |
| Toileting or elimination | 2.62 | 2.14 | 3.21 | <0.001 |
| Mental health | 5.35 | 4.85 | 5.91 | <0.001 |
| Skin, hair, or beauty | 1.96 | 1.60 | 2.39 | <0.001 |
| Sleep | 3.07 | 2.73 | 3.45 | <0.001 |
| Eyes | 2.01 | 1.74 | 2.31 | <0.001 |
| Nose | 1.54 | 1.09 | 2.17 | 0.014 |
| Ears | 2.35 | 1.74 | 3.18 | <0.001 |
| Other | 2.58 | 2.22 | 3.00 | <0.001 |

\*Adjusted for sex, age, marital status, equivalent income, education, company size, job type, frequency of working from home, number of days worked per week, presence of a policy by the employer requesting that employees refrain from attending work while ill, cumulative infection rate for COVID-19 in the region of residence, and treatment status.
OR: odds ratio, CI: confidence interval

The predictive margins of sickness presenteeism days for each treatment status and symptom are shown in Table 4 and Figure 2. When the analysis was performed based on the three treatment statuses, irrespective of symptom, the predictive margin of sickness presenteeism days among workers who experienced interrupted medical care was 6.6 days (standard error [SE]=0.25), while that among workers who did not require routine medical care was 1.4 days (SE=0.03). When the analysis was performed based on the 36 treatment-symptom groups (3 treatment statuses and 12 symptoms), the largest predictive margin of sickness presenteeism days was observed for mental health symptoms AND interrupted medical care (predictive margin: 9.9 days, SE=0.38). The simple effect comparisons test, which included the interaction term between treatment status and symptoms, showed that there were significant differences between workers with the same symptoms who did and did not require routine medical care, and between workers with the same symptoms who experienced interruption to medical care and who did not require routine medical care. For example, the fraction of sickness presenteeism days significantly differed between those with mental health symptoms who used medical care and those with mental health symptoms who did not require routine medical care (*p*<0.001).

**Table 4.** Predictive margins of sickness presenteeism days for each symptom and comparison between each treatment status

|  | Workers who did not require<br>routine medical care<br>(n=17,526) |  |  |  | Workers who used medical care<br>(n=8,451) |  |  |  |  | Workers who experienced interrupted<br>medical care<br>(n=1,059) |  |  |  |  |
| --- | --- | --- | --- | --- | --- | --- | --- | --- | --- | --- | --- | --- | --- | --- |
|  | n | % | Predictive<br>margins* | Standard<br>error | n | % | Predictive<br>margins* | Standard<br>error | *p* † | n | % | Predictive<br>margins* | Standard<br>error | *p* † |
| Total | 17526 | 100 | 1.4 | 0.03 | 8451 | 100 | 2.2 | 0.05 | <0.001 | 1059 | 100 | 6.6 | 0.25 | <0.001 |
| Main symptom |  |  |  |  |  |  |  |  |  |  |  |  |  |  |
| No problem | 10938 | 62.4 | 0.8 | 0.03 | 3642 | 43.1 | 0.9 | 0.03 | 1.000 | 160 | 15.1 | 2.6 | 0.15 | <0.001 |
| Pain | 849 | 4.8 | 2.0 | 0.10 | 842 | 10.0 | 2.5 | 0.12 | <0.001 | 144 | 13.6 | 6.1 | 0.33 | <0.001 |
| Movement | 481 | 2.7 | 2.3 | 0.16 | 344 | 4.1 | 2.8 | 0.20 | 1.000 | 54 | 5.1 | 6.5 | 0.44 | 0.428 |
| Tightness, loss of<br>energy, appetite,<br>fever, dizziness, or<br>feeling poor | 715 | 4.1 | 3.4 | 0.15 | 583 | 6.9 | 4.1 | 0.17 | 0.027 | 133 | 12.6 | 8.9 | 0.39 | <0.001 |
| Toileting or<br>elimination | 263 | 1.5 | 2.0 | 0.18 | 201 | 2.4 | 2.5 | 0.23 | 1.000 | 40 | 3.8 | 5.5 | 0.50 | <0.001 |
| Mental health | 1143 | 6.5 | 3.8 | 0.14 | 909 | 10.8 | 4.7 | 0.17 | <0.001 | 219 | 20.7 | 9.9 | 0.38 | <0.001 |
| Skin, hair, or beauty | 389 | 2.2 | 1.5 | 0.14 | 160 | 1.9 | 1.9 | 0.18 | 0.006 | 29 | 2.7 | 4.9 | 0.44 | 0.513 |
| Sleep | 997 | 5.7 | 2.3 | 0.11 | 637 | 7.5 | 2.8 | 0.14 | 0.127 | 116 | 11.0 | 6.5 | 0.35 | <0.001 |
| Eyes | 795 | 4.5 | 1.5 | 0.09 | 512 | 6.1 | 1.8 | 0.11 | 0.319 | 65 | 6.1 | 4.3 | 0.30 | <0.001 |
| Nose | 98 | 0.6 | 1.2 | 0.20 | 71 | 0.8 | 1.5 | 0.25 | 1.000 | 9 | 0.9 | 3.4 | 0.54 | 1.000 |
| Ears | 80 | 0.5 | 1.7 | 0.25 | 84 | 1.0 | 2.2 | 0.30 | 1.000 | 12 | 1.1 | 5.7 | 0.73 | 0.056 |
| Other | 778 | 4.4 | 2.0 | 0.13 | 466 | 5.5 | 2.5 | 0.17 | 0.031 | 78 | 7.4 | 6.3 | 0.41 | <0.001 |

\*predictive margins: mean predicted number of sickness presenteeism days for each symptom and treatment status using the model in Table 2 and 3 with substitution of measured values for other covariates (adjusted using the Bonferroni method); calculated using the formula: mean predicted fraction × 30 (days)
<sup>†</sup> $p$ -value for simple main effects for each treatment status compared to workers who did not require routine medical care using a model that included the interaction term for treatment status and main symptoms (adjusted using the Bonferroni method)
%: proportion of the total number of respondents for each treatment status

**Figure 2.**
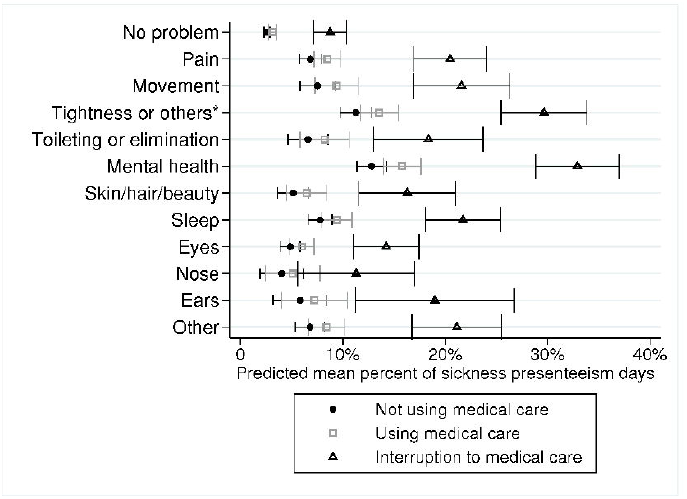
Predictive margins with 95% confidence intervals for each treatment status and symptom *others include loss of energy, appetite, fever, dizziness, or feeling poor †Error bars indicate 95% confidence intervals

## Discussion

This study demonstrated an association between treatment interruption and sickness presenteeism among Japanese workers during the country’s third wave of the COVID-19 pandemic. Compared to workers who did not require routine medical care, workers who had diseases that required routine medical care reported more days of sickness presenteeism, and those who experienced interrupted medical care reported even more such days. Furthermore, our findings revealed that there may be some symptoms that are more likely to lead to sickness presenteeism.

We found that workers who experienced interrupted medical care had increased sickness presenteeism. This is because appropriate treatment can improve work function and productivity by improving workers’ health and subjective symptoms^10,28^. Employees who experience interrupted treatment for chronic diseases may be forced to return to work due to fear of being laid off, depending on the financial situation of their workplace during the pandemic. It is important to continue regular treatments to manage disease and maintain health^29^ to prevent inappropriate presence at work while ill.

We found that the proportion of sickness presenteeism days varies by the type of symptoms experienced by workers. While ORs among workers with symptoms related to mental health problems, loss of energy/fever, body movement, and sleep were high, those among workers with nose and skin, hair, beauty symptoms were moderate. Sickness presenteeism is the result of a worker’s choice to attend work despite being unwell rather than being absent from work. Many previous studies conducted mainly in Europe have evaluated “sickness presenteeism” as a health behavior based on whether or not workers “worked one or more days in a certain period of time with a health condition for which they think they really should be absent”^2,30^. This is in contrast to studies of “presenteeism” in North America, where the concept is evaluated as productivity loss due to illness or a health condition using the Work Limitations Questionnaire or other tools^10,31^. One reason as to why sickness presenteeism is more likely to be reported by workers experiencing mental health problems may be that these symptoms are severe enough to cause “health conditions for which they think they really should be absent.” In contrast, physical symptoms such as those of allergic rhinitis may not be considered “health conditions for which they think they really should be absent” regardless of severity. If workers themselves do not consider their symptoms to be “health conditions for which they think they really should be absent,” they may not report experiencing sickness presenteeism. However, there may be cases in which workers do not deem it necessary to be absent from work, despite having a symptom that reduces productivity. Allergies with nose-related symptoms are one example that causes loss of productivity due to presenteeism^11,32^. Workers may not link these symptoms to sickness presenteeism due to differences in interpretation of “health conditions that require absence from work.” This is an important point when evaluating “sickness presenteeism” as a health behavior.

We also found that the impact of continuing treatment on the prevention of sickness presenteeism varied by symptom. Sickness presenteeism was more frequent in workers with treatment interruption and symptoms related to pain, tightness and loss of energy, toileting and elimination, mental health, sleep, and eyes than those who did not require routine medical care. Among those who reported toileting and elimination, sleep, and eye symptoms, there was no difference in sickness presenteeism between workers who used medical care and those who did not require routine medical care. This suggests the importance of continuing necessary routine medical care for preventing sickness presenteeism due to these symptoms. For those with pain, tightness and loss of energy, and mental health symptoms, sickness presenteeism remained high even with continued treatment, indicating the need to identify appropriate treatment and manage one’s daily health condition in addition to continuing treatment. Meanwhile, the prevalence of sickness presenteeism was comparable among treatment statuses for participants with movement and mobility, nose, and ear symptoms. This may be because individuals may not consider these symptoms sufficiently adverse to engage in sickness presenteeism, or may experience chronic symptoms for which support and adaptive behaviors have already been put into place such as movement symptoms. The impact of continuing treatment on sickness presenteeism may be related to whether an individual considers their symptoms to be sufficiently adverse to require an absence from work, or whether or not the symptoms can be improved with treatment.

There are several limitations to this study. First, since this study is a survey of Internet monitors, limitations regarding selection bias and generalizability are unavoidable. To reduce potential bias, sampling and recruitment were conducted according to occupation and sex in each region and the COVID-19 infection rate. To understand the characteristics of the target population of this survey, we compared the results with those of national surveys and occupational surveys using various batteries^23^. Second, we did not obtain detailed information related to treatment interruptions, including the type of disease, duration, and reasons for interruption. We were thus unable to determine whether the reason for interruption to treatment was due to patient-related reasons (e.g., economic situation and anxiety) or hospital-related reasons (e.g., schedule adjustment). Additionally, the questions we used to identify exposure factors were related to the presence or absence of diseases that require hospital visits and use of medical care. However, sickness presenteeism is not just related to disease and medical status, but also a wide range of health conditions or concerns. Third, interruptions to treatment may be the result of better disease control and improved health. It is unclear how these factors would affect the occurrence of sickness presenteeism. Fourth, because this study examined sickness presenteeism among workers during the third wave of the COVID-19 pandemic in Japan, caution is needed when interpreting the results. Further, causality is unclear. For example, because most workers were told to remain home if they were experiencing COVID-like symptoms during the pandemic, sickness presenteeism may have been reduced during this period. However, it is unclear how such instructions affected overall sickness presenteeism, including that associated with symptoms unrelated to COVID-19. Finally, we did not consider all possible confounders affecting sickness presenteeism because we did not obtain information on some confounders, such as job insecurity, annual leave rights, and the culture around employment and sick leave in each company.

### Conclusion

Interruption to medical care was associated with the occurrence of sickness presenteeism during Japan’s third wave of the COVID-19 pandemic. While the occurrence of sickness presenteeism largely differed according to symptoms, it may be possible that there were important cases of sickness presenteeism that were undetectable using the questionnaire. This study demonstrates the importance of maintaining one’s health condition and continuing necessary treatment even during an infectious disease pandemic.

## Data Availability

Data not available due to ethical restrictions

## Acknowledgements

This study was supported and partly funded by research grants from the University of Occupational and Environmental Health, Japan (no grant number); the Japanese Ministry of Health, Labour and Welfare (grant numbers H30-josei-ippan-002, H30-roudou-ippan-007, 19JA1004, 20JA1006, 210301-1, and 20HB1004); Anshin Zaidan (no grant number); the Collabo-Health Study Group (no grant number); and Hitachi Systems, Ltd. (no grant number); and scholarship donations from Chugai Pharmaceutical Co., Ltd. (no grant number).

The current members of the CORoNaWork Project, in alphabetical order, are as follows: Dr. Yoshihisa Fujino (present chairperson of the study group), Dr. Akira Ogami, Dr. Arisa Harada, Dr. Ayako Hino, Dr. Hajime Ando, Dr. Hisashi Eguchi, Dr. Kazunori Ikegami, Dr. Kei Tokutsu, Dr. Keiji Muramatsu, Dr. Koji Mori, Dr. Kosuke Mafune, Dr. Kyoko Kitagawa, Dr. Masako Nagata, Dr. Mayumi Tsuji, Ms. Ning Liu, Dr. Rie Tanaka, Dr. Ryutaro Matsugaki, Dr. Seiichiro Tateishi, Dr. Shinya Matsuda, Dr. Tomohiro Ishimaru, and Dr. Tomohisa Nagata. All members are affiliated with the University of Occupational and Environmental Health, Japan.

## Disclosure

### Ethics Approval

This study was approved by the ethics committee of the University of Occupational and Environmental Health, Japan (reference No. R2-079 and R3-006).

### Informed Consent

Informed consent was obtained using a form on the survey website.

### Registry and the Registration No. of the study/Trial

N/A

### Animal Studies

N/A

